# The evaluation of the quality of life in survivors of critical illness after discharge from Intensive Care Unit (ICU): A prospective cohort study

**DOI:** 10.1101/2023.04.24.23289053

**Authors:** Dimitrios Papageorgiou, Christos Triantafyllou, Areti Stavropoulou, Martha Kelesi, Evridiki Kaba, Konstantinos Evgenikos, Athanasia Liveri, Theodora Stroumpouki

## Abstract

**Introduction:** Admission to the Intensive Care Unit (ICU) constitutes a substantial psychophysical burden for both patients and their relatives. Individuals who are critically ill and receive care in the ICU frequently exhibit numerous physical and mental issues stemming from the primary ailment, its complications, and subsequent treatments. This study aimed to assess the quality of life (QoL) for ICU patients in the first and third month after ICU discharge and identify any issues arising from their ICU hospitalization.

**Material and methods:** This was a prospective cohort study. The study participants were adult medical or surgical patients admitted to Intensive Care Units (ICUs) in three public hospitals in the region of Attica in Greece, from August 2020 to December 2021. The short form (SF)-36 was used to measure QoL. Data collection was performed through telephone interviews during the first and third month after ICU discharge.

**Results:** The study included 43 patients, 34 men and nine women. The mean age was 59.63±13.06 years. The average value of the two main categories in the 1st month was: Physical health: 53.72 ± 15.92, and Mental health: 69.03 ± 26.02, while in the 3^rd^ month, it was 62.42 ± 20.45 and 72.81 ± 16.47, respectively. The duration of mechanical ventilation, high-flow oxygen therapy, and spontaneous breathing in days seemed to be correlated with the “Physical Functioning”, “Pain”, and “Limitation of the role due to Physical health” subscales of SF-36, respectively (p-value <0.05). The total length of hospitalization seemed to have a statistically significant negative correlation with “Physical Function” and “Physical Health” subscales (p-value<0.05).

**Conclusions:** An improvement in patients’ QoL was demonstrated three months after discharge from the ICU. Factors such as gender, cause of ICU admission, occurrence or non-occurrence of cardiac arrest, the performance of a tracheostomy, and type of ventilation support should always be considered when assessing the quality of life after ICU hospitalization.

## Introduction

Admission to the Intensive Care Unit (ICU) constitutes a substantial physical and psychological burden for both patients and their relatives *(1)*. Individuals who are critically ill and receive care in the ICU frequently exhibit numerous physical and mental issues stemming from the primary ailment, its complications, and subsequent treatments *(1)*. Nevertheless, despite many ICU patients recuperating from severe conditions, they might still face persistent issues at post-discharge, such as dyspnea, debilitation, anxiety, depression, sleep disruption, difficulty concentrating, post-traumatic stress, and pain *(2)*. These issues can impair daily activities, work or educational pursuits, interpersonal connections, social interactions, and the financial stability of the patient and/or their family *(2)*. As a result, ICU hospitalization represents a distressing event with a considerable influence on the patients’ overall well-being.

Historically, the primary objective of ICUs was to help patients manage their critical health conditions and transition back to the general nursing ward for ongoing care *(3)*. In those days, the mere transfer of a patient to the nursing ward, irrespective of the often imminent death, was considered a success of ICU treatment *(4)*. However, as medicine and nursing science have advanced over time, these circumstances have transformed, altering the ultimate aim of care administered in ICUs *(5)*. Presently, the focus is on providing the highest quality of intensive care, enabling patients not only to survive but also to return to their pre-illness state with minimal complications and disabilities *(5,6)*. This progress and the necessity for comprehensive care have prompted questions about the quality of life for patients following ICU and hospital discharge *(7)*. In essence, there emerged a need to track these patients’ subsequent lives in the community, document the novel challenges they face in their daily routines, and determine whether these challenges enable the patients to be functional *(8)*. These challenges encompass any acquired disabilities, life support measures (such as mechanical ventilation or feeding devices), as well as psychosomatic consequences like fatigue, stress, and post-traumatic stress disorder (PTSD) resulting from hospitalization *(9)*. Naturally, monitoring a patient’s progress during the post-discharge period should also take into account their immediate surroundings, including their family *(10)*. The family environment, which encompasses the patient, must adapt to the new circumstances and may potentially assume the role of a caregiver or supporter, significantly contributing to the patient’s recuperation *(11)*.

Research at an international level has indicated that the quality of life (QoL) for patients who survive ICU hospitalization is still uncertain *(3,12,13)*. In Greece, there have been limited efforts to evaluate the impact of ICU hospitalization on patients’ quality of life (14–16).

Consequently, the study’s objective was to assess the QoL for ICU patients in the first and third month after ICU discharge and identify any issues arising from their ICU hospitalization.

## Material and Methods

### Study Design and Participants

This was a prospective cohort study conducted in three public hospitals in the Attica region of Greece the study participants were adult medical or surgical patients admitted to Intensive Care Units (ICUs) from August 2020 to December 2021. Inclusion criteria for the participation in the study were (1) patients needed to be over 16 years old; (2) had received mechanical ventilation for at least 1 hour; (3) were expected to survive ICU care after stabilization as determined by clinicians’ assessments; and (4) had satisfactory Greek reading and writing skills. The study did not include patients with an ICU stay shorter than 24 hours or a diagnosed mental illness. Convenience sampling was employed, and patients were enrolled either during their ICU stay or after being transferred out of the ICU but before hospital discharge.

### Data collection

For all included patients, the following data were collected: age, gender, educational background, marital status, the reason for admission, type of tracheotomy (Surgical procedure or percutaneous technique), duration of mechanical ventilation, length of stay in ICU, length of hospitalization, patient outcome (discharged or deceased), and any readmission within 30 days from hospital discharge.

The short form (SF)-36 was used to measure QoL. SF-36 *(17,18)* is a 36-item patient-reported outcome questionnaire that covers eight health domains: physical functioning (10 items), pain (2 items), role limitations due to physical health (4 items), role limitations due to personal or emotional problems (4 items), emotional well-being (5 items), social functioning (2 items), energy/fatigue (4 items), and general health (5 items). Scores for each domain range from 0 to 100, with a higher score defining a more favorable health state. SF-36 has demonstrated reliability, validity, and responsiveness in the post-ICU population *(17)* and is one of the most common instruments used for assessing health status in this patient cohort *(19,20)*.

Three research nurses assessed the quality of life using a telephone assessment of SF-36. The three research nurses piloted the interview technique in 20 patient telephone interviews. The patients completed the SF-36 questionnaires one month and three months after ICU discharge.

### Ethical issues

Regarding the ethics of this study, the ethical principles described by the Belmont Report (1976) on the rights of research subjects were applied *(21)*. In addition, the principles of (1) benefit and no harm, (2) respect for human dignity, and (3) fairness were considered at all stages of the study. The study was approved by the hospitals’ review boards (Ref No 83/26-2-2020, 37/10-2-2020, and 27/25-2-2020). Data collection and analysis were conducted after obtaining informed, written consent from all patients or their relatives during ICU care and from all patients once they regained competency. The patients’ personal data and the hospitals’ names remained anonymous at all stages of the study.

### Statistical analysis

The Shapiro-Wilk statistical test was used to check the normality of the data. Mean (M), Standard Deviation (SD), and range were used to describe the quantitative variables, while absolute (n) and relative frequencies (%) were used to describe the qualitative variables. The reliability of the SF-36 was tested using Cronbach’s Alpha reliability coefficient. Each scale showed satisfactory reliability (index values > 0.7). To investigate the correlation between two quantitative variables, Pearson’s correlation coefficient was used parametrically, and Spearman’s correlation coefficient was used non-parametrically. The comparison between a quantitative variable and a qualitative variable with two levels was performed using a parametric Student’s t-test and non-parametric Mann-Whitney U test. Finally, to compare two dependent samples, the Paired-Samples T-Test was used parametrically, and Wilcoxon test was used non-parametrically. All tests were performed at a 0.05 level of significance. The statistical package for Social Sciences (SPSS) ver.24 was used for data analysis and processing.

## Results

### Characteristics of Patients

The study included 43 patients (34 men: 79.1%, and nine women: 20.9%). The mean age was 59.63±13.06 years. The demographic characteristics of the participants are presented in **Table 1**.

**Table 1.** Demographic characteristics of participants

|  | **n** | **%** |
| --- | --- | --- |
| Sex |  |  |
| Male | 34 | 79.1 |
| Female | 9 | 20.9 |
| Educational level |  |  |
| Primary education | 2 | 4.7 |
| Secondary education | 19 | 44.2 |
| Higher education | 22 | 51.2 |
| Postgraduate studies |  |  |
| Yes | 2 | 4.7 |
| No | 41 | 95.3 |
| Occupation |  |  |
| Civil servant | 3 | 7.0 |
| Private employee | 15 | 34.9 |
| Freelancer | 8 | 18.6 |
| Unemployed | 1 | 2.3 |
| Retired | 16 | 37.2 |
| Marital status |  |  |
| Married | 33 | 76.7 |
| Unmarried | 6 | 14.0 |
| Divorced | 1 | 2.3 |
| Widower | 3 | 7.0 |
| No of children |  |  |
| 0 | 8 | 11.6 |
| 1 | 11 | 51.2 |
| 2 | 17 | 20.9 |
| 3 | 5 | 11.6 |
| > 3 | 2 | 4.7 |
| How many people stay with the patient? |  |  |
| 0 | 5 | 11.6 |
| 1 | 22 | 51.6 |
| 2 | 9 | 20.9 |
| 3 | 5 | 11.6 |
| > 3 | 2 | 4.7 |
| Social Security |  |  |
| Uninsured | 3 | 7.0 |
| Public | 36 | 83.8 |
| Private | 4 | 9.3 |
| Monthly income |  |  |
| 0-1000 | 11 | 25.6 |
| 1001-2000 | 28 | 65.1 |
| 2001-3000 | 3 | 7.0 |
| >3000 | 1 | 2.3 |
|  | **Mean ±SD** | **Range** |
| Patient age in years | 59,63±13,06 | 25-81 |

Regarding the reasons for the admission of patients to the ICU, 74.4% (n=32) had respiratory problems, of which 12 (27.9%) also had COVID-19. Cardiac arrest was suffered by 16.3% (n=7), reintubation was performed in 16.3% (n=7), and tracheostomy was performed in 11.6% (n=5) of the patients. 90.7% (n=39) of patients were transferred to an inpatient department from the ICU, and only 4.7% (n=2) were readmitted within 30 days to the ICU. The total length of hospitalization was 30.16±33.91 days.

### Comparison between the first and third month of follow-up

**Table 2** shows the differences observed in the SF-36 subscale scores between the 1^st^ and 3^rd^ month of follow-up. The subscales “Role limitation due to Emotional Problems”, “Mental Health”, and “Pain” did not appear to be statistically significantly different between the 1^ST^ and 3^rd^ month of follow-up (p-value>0.05). The other subscales showed a statistically significant difference between the 1^st^ and 3^rd^ month of follow-up (p-value<0.05) with an almost overall increase in scores except for the “General Health” dimension.

**Table 2.**
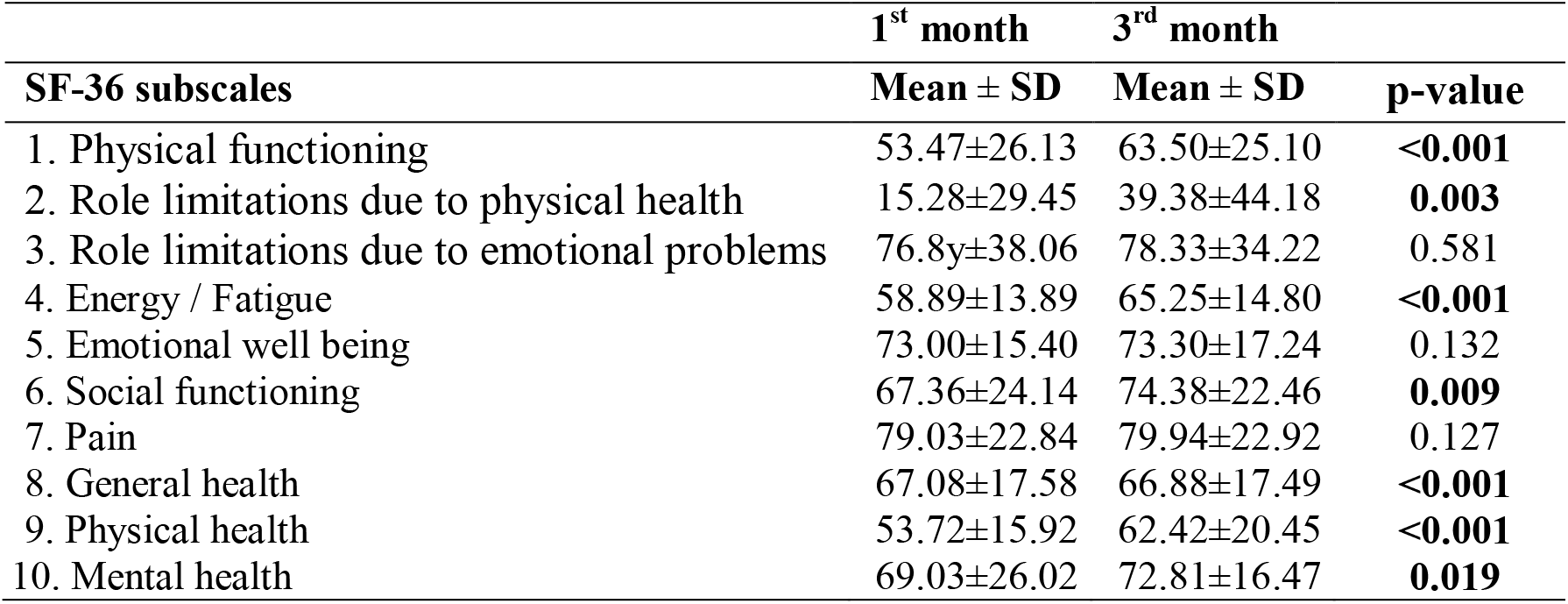
Comparisons of SF-36 subscales at the 1^st^ and 3^rd^ month of the follow-up

|  | **1<sup>st</sup> month** | **3<sup>rd</sup> month** |  |
| --- | --- | --- | --- |
| **SF-36 subscales** | **Mean ± SD** | **Mean ± SD** | **p-value** |
| 1. Physical functioning | 53.47±26.13 | 63.50±25.10 | **<0.001** |
| 2. Role limitations due to physical health | 15.28±29.45 | 39.38±44.18 | **0.003** |
| 3. Role limitations due to emotional problems | 76.8y±38.06 | 78.33±34.22 | 0.581 |
| 4. Energy / Fatigue | 58.89±13.89 | 65.25±14.80 | **<0.001** |
| 5. Emotional well being | 73.00±15.40 | 73.30±17.24 | 0.132 |
| 6. Social functioning | 67.36±24.14 | 74.38±22.46 | **0.009** |
| 7. Pain | 79.03±22.84 | 79.94±22.92 | 0.127 |
| 8. General health | 67.08±17.58 | 66.88±17.49 | **<0.001** |
| 9. Physical health | 53.72±15.92 | 62.42±20.45 | **<0.001** |
| 10. Mental health | 69.03±26.02 | 72.81±16.47 | **0.019** |

### Comparison between the demographic characteristics of patients in the 1^st^ and 3^rd^ month of the follow-up

During the 1st month follow-up, the comparison of participants’ demographic characteristics and SF-36 subscales showed statistically significant differences between the cause of ICU admission (COVID-19 or otherwise) and the “Social Functioning” subscale (p-value<0.05), between the occurrence or non-occurrence of cardiac arrest and the “Energy/Fatigue” subscale (p-value<0.05), and between the presence or absence of tracheostomy and the “Physical Health” subscale (p-value<0.05). Patients with COVID-19 seemed to engage in social activities more effectively, without being hindered by the physical and emotional issues stemming from their illness, as compared to patients admitted for other reasons. Moreover, patients who experienced cardiac arrest appeared to feel exhausted occasionally due to their health condition, in contrast to those who did not. Additionally, patients who underwent tracheostomy exhibited significant self-care limitations, frequently experienced fatigue, and assessed their health as poor compared to patients who did not have a tracheostomy (Table 3).

**Table 3.**
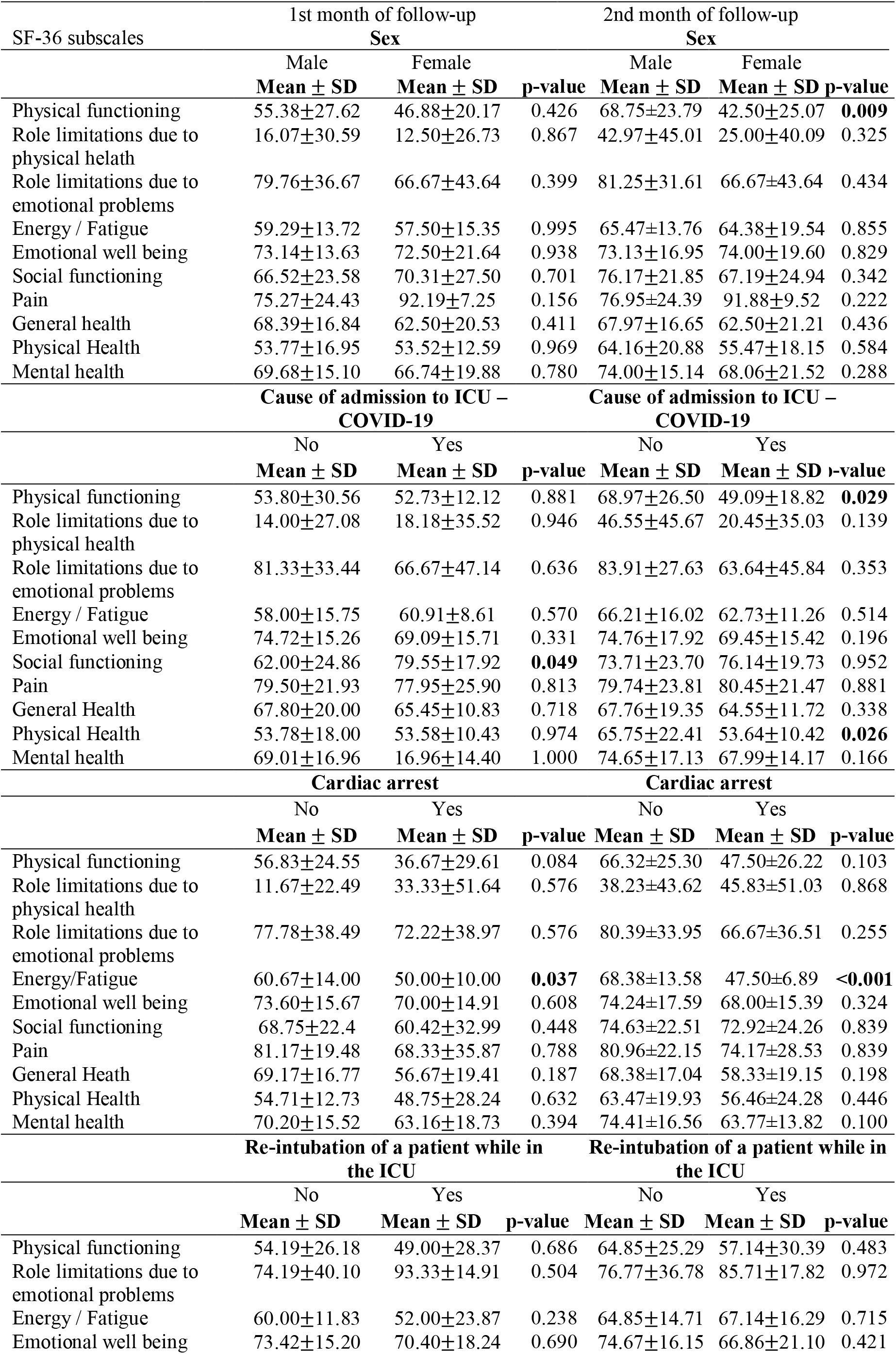
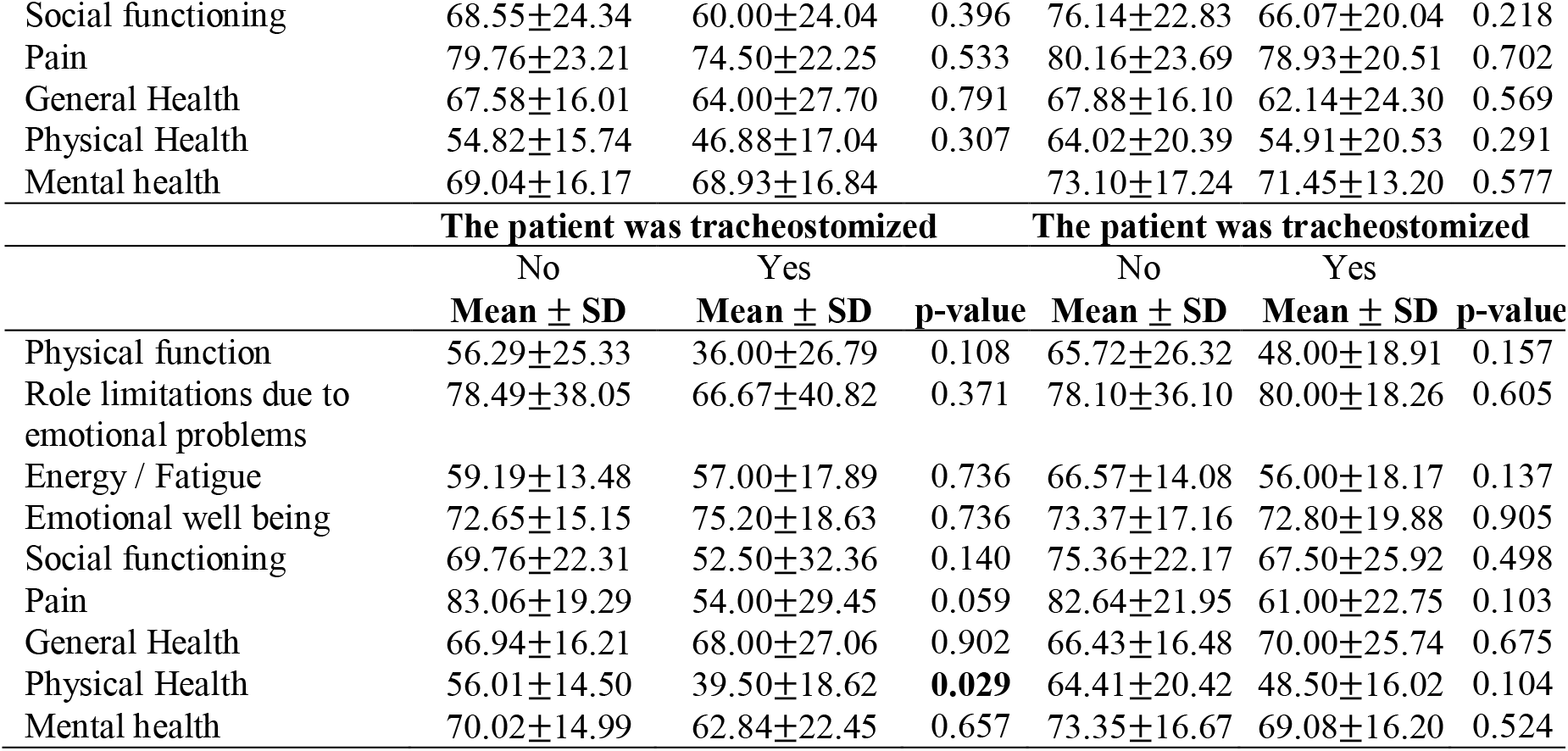
Comparisons of SF-36 scales for the 1st and 3rd month of follow-up with participants’ demographic characteristics (n=43)

Regarding the 3^rd^ month of follow-up, a statistically significant difference was noted between the patients’ gender and the “Physical Function” subscale (p-value<0.05), between the cause of ICU admission (COVID-19 or other) and both the “Physical Function” and “Physical Health” subscales (p-value<0.05), as well as between the occurrence or non-occurrence of cardiac arrest and the “Energy/Fatigue” subscale (p-value<0.05). Female patients appeared to be more constrained in various physical functions, such as dressing and bathing independently, compared to male patients. Patients with COVID-19 seemed to experience greater limitations in several physical functions, like dressing and bathing alone and perceived their health to be worse than those admitted for other reasons. Lastly, patients who experienced cardiac arrest appeared to feel fatigued and exhausted at times due to their health condition, in contrast to those who did not have a cardiac arrest (Table 3).

During the 1st month follow-up, the length of ICU hospitalization (in days) appeared to have a slight negative correlation with the “Physical Function” and “Energy/Fatigue” subscales. The duration of mechanical ventilation (in days) seemed to be slightly negatively correlated with the “Physical Function,” “Pain,” and “Physical Health” scales. The duration of high-flow oxygen therapy (in days) displayed a very slight positive correlation with the “Pain” scale. The length of hospitalization with spontaneous breathing (in days) demonstrated slight negative correlations with the “Physical Function,” “Energy/Fatigue (or Vitality),” and “Mental Health” subscales. The total length of hospitalization (in days) appeared to have a slight negative correlation with the “Physical Function” and “Energy/Fatigue” subscales.

In contrast, during the 3^rd^ month follow-up, the duration of mechanical ventilation (in days) appeared to have a slight negative correlation with the “Pain” subscale. The duration of high-flow oxygen therapy (in days) displayed a slight positive correlation with the “Pain” subscale. The length of hospitalization with spontaneous breathing (in days) demonstrated a slight negative correlation with the “Physical Function” subscale. The total length of hospitalization (in days) seemed to have a slight negative correlation with both the “Physical Function” and “Physical Health” subscales.

Patients’ age did not appear to correlate with any SF-36 subscale in either the 1^st^ or 3^rd^ month of follow-up.

## Discussion

The current study revealed that the quality of life for patients who were hospitalized in the ICU improved in the 3rd month of follow-up compared to the 1st month. Specifically, the subscales “Physical Function,” “Role Limitation due to Physical Health,” “Energy/Fatigue,” “Social Function,” “General Health,” “Physical Health,” and “Mental Health” demonstrated higher scores during the 3rd month of patient follow-up. This suggests a slight improvement in health, highlighting the benefits of the treatment received by patients within the ICU. This finding is consistent with that of a previous study *(14)*. Specifically, in a study conducted in Greece, which aimed to assess the changes in health-related quality of life (HRQoL) in patients discharged from the ICU using the “Quality of Life-Spanish (QOL-SP)”, it was found that the mean quality of life score of the patients increased from 2.9 ± 4.8 (out of a maximum of 25 points) upon ICU admission to 7.0 ± 7.2 points at six months after discharge, and then decreased to 5.6 ± 6.9 points at 18 months (p-value<0.001) *(14)*.

Moreover, a systematic review encompassing 48 studies with a total of 11,927 patients reported that quality of life improved in the first year after discharge in four domains: physical function, physical role, vitality, and social function, with no significant improvement observed thereafter *(22)*. However, these domains were also the least likely to return to population norms, as they were more deeply impacted by critical illness *(22)*. The critical illness appears to affect psychological health less than physical health, and the former may experience less improvement following hospital discharge *(22)*. Interventions aimed at enhancing health after critical illness may be more effective for physical health than psychological health *(22)*.

In this study, we found that age is not associated with the quality of life following discharge, which is consistent with the findings of two previous studies *(13,23)*. In a study conducted in France *(13)*, for patients older than 70, scores were not different from those of younger patients, whatever the underlying condition. Another study conducted by Orwelious et al. *(23)*, it was found that age did not have any impact on health-related quality of life (p-value=0.084). However, this result contrasts with the findings of two other previous studies *(14,24)*. In a Greek study *(14)*, it was found that age had a positive association with quality of life at 18 months after ICU discharge (B = 0.129, p-value<0.001). In another study conducted by Vogel et al. *(24)*, which aimed to describe and analyze factors associated with Health-Related Quality of Life (HRQoL) after discharge from a general surgical ICU using the SF-36, it was found that patients aged 65-74 years estimated their HRQoL lower than both younger and older patients in two domains: Physical Functioning (p-value = 0.00) and Mental Health (p-value = 0.04).

In the 3^rd^ month of follow-up, the female gender exhibited a lower quality of life, which is consistent with three previous studies *(14,24,25)*. Vogel et al. *(24)* found that the female gender was associated with lower HRQoL in three domains of SF-36; Bodily Pain (p-value = 0.03), Emotional Role (p-value = 0.04), and Mental Health (p-value= 0.01). In a Portuguese study *(25)*, which aim was to assess HRQOL and independence in activities of daily living (ADL) six months after discharge from an ICU, and to study its determinants, it was found that women had significantly lower scores for bodily pain, general health perception, vitality, and social functioning than men. Fildissis et al. *(14)* found that the male gender had a positive association with quality of life at 18 months after ICU discharge (B = 3.934, p-value=0.002).

Another interesting finding of our study was that during the 1^st^ and 3^rd^ month follow-up, statistically significant differences were observed between the cause of ICU admission (COVID-19 or otherwise) and the “Social Functioning” subscale, as well as between the cause of ICU admission (COVID-19 or other) and both the “Physical Function” and “Physical Health” subscales, respectively. A similar study involving COVID-19 patients has not been conducted. However, other studies suggest that car accident victims who survived serious injuries experienced significant levels of disability with subsequent reduced quality of life *(26,27)*.

Another interesting finding of our study was that patients who underwent tracheostomy appeared to have significant limitations in their self-care, often felt fatigued, and rated their quality of life as poor compared to patients who did not undergo tracheostomy. This finding is in contrast with this of a previous study *(28)*. Specifically, in an Italian study that aims to investigate post-discharge survival and QoL after tracheostomy for Acute respiratory failure (ARF) in patients with amyotrophic lateral sclerosis (ALS) using Life Satisfaction Index (LSI-11), it was found that the mean (SD) cumulative score on the LSI-11 was 9.3 (3.6; range, 0-22; higher values indicating better QoL), similar to that obtained from a control group consisting of individuals with ALS who had not received tracheostomy (9.3 ± 4.3) and to that reported for persons in the general population. This may be due to the fact that the variation in patient-perceived life satisfaction may not solely be linked to physical function; it could also be associated with sociodemographic factors such as income, social life, and education *(29)*.

In the present study, we found that the length of ICU hospitalization is negatively and weakly correlated with the “Physical Function” and “Physical Health” of SF-36 subscales. This finding is in line with this of a previous study *(15)*. In a study conducted by Vrettou et al. *(15)*, which aimed to investigate the quality of life using the WHOQOL-Bref Questionnaire and the correlations of clinical and psychological parameters with the quality of life scores in survivors of critical illness one year after discharge from intensive care, it was found that the ICU stay duration correlated negatively with the physical (Pearson’s r = -0.19, p-value = 0.04) and the social relationship (Pearson’s r = -0.19, p-value = 0.04) domains of WHOQOL-Bref Questionnaire.

Finally, one more interesting finding was that the duration of mechanical ventilation in days appears to be negatively and slightly correlated with the “Pain” subscale. In a systematic published in 2010 *(12)*, which aim was to evaluate the quality of life at least 12 months after discharge from the intensive care unit of adult critically ill patients and to give an overview of factors influencing the quality of life, it was found that the worst reductions in quality of life were seen in cases of prolonged mechanical ventilation.

Our study has some limitations that should be acknowledged. First of all, the sample size was small, as patients were recruited from general adult ICUs of only three Greek hospitals. Thus, the study sample may not be fully representative, and accordingly, the generalization of the present study’s findings should be interpreted with caution. Another important limitation is that results are exclusively based on self-reported data; participants may provide an answer that is widely accepted rather than reflecting their true beliefs, a phenomenon called socially desirable responding (SDR), leading to response bias *(30)*. Lastly, the assessment of QoL in this study was restricted to 3 months after discharge, meaning that the patients who participated were still in the recovery phase. A longer study period is recommended to illustrate more explicitly the ICU patients’ condition after discharge.

## Conclusions

In conclusion, the benefits of ICU treatment were demonstrated as there was a slight improvement in the QoL of the patients in the third month after ICU discharge. Factors such as gender, cause of ICU admission, occurrence or non-occurrence of cardiac arrest, the performance of a tracheostomy, and type of ventilation support should always be considered when assessing the quality of life after ICU hospitalization.

QoL assessments after ICU discharge should be conducted more frequently, as they can potentially provide insights into the long-term outcomes of ICU survivors. Additionally, these studies may help identify the main challenges that patients face after discharge, thereby enabling the design of specific follow-up programs based on their needs.

Finally, further studies are needed to improve QoL assessment, allowing comparisons between ICUs and enhancing clinicians’ skills and knowledge of patients who experience problems after discharge due to ICU hospitalization. Although the nursing staff can perform certain aspects of early intervention in ICUs, the high percentage of patients underscores the need for access to specialist services. This preliminary validation study suggests that the SF-36 Health Survey Questionnaire could be reliably used as a screening instrument at six months, 1 year, and every year post-discharge from ICU to identify patients needing referral to such services. This is particularly useful as it aligns well with the timing of ICU follow-up clinics being established in the rest of Europe and recommendations for the timing of early intervention.

## Data Availability

All data produced in the present study are available upon reasonable request to the authors.

